# Neuroinflammation, Depressive Affect, and Amyloid Burden in Alzheimer’s Disease: Insights from the Kynurenine Pathway

**DOI:** 10.1101/2020.08.11.20172940

**Authors:** Auriel A. Willette, Colleen Pappas, Nathan Hoth, Qian Wang, Brandon Klinedinst, Sara A. Willette, Brittany Larsen, Amy Pollpeter, Tianqi Li, Scott Le, Jonathan P. Mochel, Karin Allenspach, Robert Dantzer, and for the Alzheimer’s disease Neuroimaging Initiative

**Author notes:** **Address Correspondence to:** Auriel A. Willette, Ph.D., M.S. (Mr.), 1109 HNSB, 706 Morrill Rd., Ames, IA 50011.

## Abstract

**Background:** Depressive symptoms in Alzheimer’s disease (AD) predict worse cognitive and functional outcomes. Both AD and major depression are characterized by shunted tryptophan metabolism away from serotonin (5-HT) and toward the neuroinflammatory kynurenine (Kyn) pathway. The present study assessed the role of Kyn across the AD continuum in behavioral, neuroanatomical, neuropathological, and physiological outcomes.

**Methods:** In 746 participants from the Alzheimer’s Disease Neuroimaging Initiative-1 (ADNI1) cohort, serum markers of 5-HT, tryptophan, and Kyn were measured and their relationships investigated with immunologic markers, affect and functional outcomes, CSF markers of beta-amyloid (Aβ) and tau, and regional gray matter.

**Results:** A higher Kyn/Tryptophan ratio was linked to many inflammatory markers, as well as lower functional independence and memory scores. A higher Kyn/5-HT ratio showed similar associations, but also strong relationships with depressive affect and neuropsychiatric disturbance, executive dysfunction, and global cognitive decline. Further, gray matter atrophy was seen in hippocampus, anterior cingulate, and prefrontal cortices, as wel as greater amyloid and total tau deposition. Finally, using moderated-mediation, several pro-inflammatory factors partially mediated Kyn/5-HT and depressive affect scores in participants with subclinical Aβ (i.e., Aβ-), whereas such associations were fully mediated by Complement 3 in Aβ+ participants.

**Conclusions:** These findings suggest that neuroinflammatory signaling cascades may occur during AD, resulting in increased Kyn metabolism that influences the pathogenesis of depressive symptoms. Aβ and the complement system may be critical contributing factors in this process.

## Introduction

Depressive affect, characterized by anhedonia, negativity, and cognitive decline, impairs overall health and is comorbid with several inflammatory and metabolic diseases. In particular, increased neuroinflammation exacerbates mood and cognitive deficits in old age (1-3). Recent evidence supports the role of inflammation in the pathophysiology of psychiatric disorders like major depression. Clinical depression is typified by higher peripheral levels of cardinal pro-inflammatory cytokines like tumor necrosis factor-α (TNF-α) and interleukin-6 (IL-6) (4). Administration of cytokine inducers, such as lipopolysaccharide (LPS), causes physical symptoms of sickness, depression, and fatigue in both animal models and human participants (5, 6). Conversely, administration of cytokine antagonists to patients with chronic inflammatory conditions, or to depressed patients with elevated biomarkers of inflammation, attenuates symptoms of depression.

The link between inflammation and depression is especially important in the context of age-related neurological disorders such as Alzheimer’s disease (AD). Approximately 30-40% of patients with AD manifest mild to major depressive symptoms (7). These behavioral symptoms and cognitive deficits may arise due to neuroinflammation and atrophy in medial temporal and prefrontal regions (8). Indeed, neuroinflammation contributes to depressive symptoms on the Geriatric Depression Scale (GDS) in old age (12). Further, this process may be exacerbated by amyloid plaques and neurofibrillary tau tangles that typify AD (9, 10), although results have been mixed (11). Participants with lifelong major depression but not AD or other cognitive impairment also show modestly more amyloid deposition in the precuneus and frontal regions (13). Perhaps not coincidentally, these brain areas are the first to show accumulation of amyloid plaques in AD, which may induce or exacerbate neuroinflammation.

Several different processes stemming from neuroinflammation may lead to increased risk for both major depression and AD, including depletion of monoamines like dopamine and serotonin (14, 15). Depletion of serotonin can occur as a result of many mechanisms, including deficient synthesis from its amino acid precursor tryptophan, or increased reuptake of serotonin at the synaptic level. Under normal physiological conditions, tryptophan is utilized for protein synthesis, neurotransmitter formation (16), and energy production via NAD/NADPH synthesis (17). Tryptophan can be hydroxylated to form 5-hydroxytryptophan, which subsequently undergoes decarboxylation to synthesize 5-hydroxytryptamine (5-HT) or serotonin. Elevated levels of Kyn have been observed with higher BMI (18), which has also been tied to neuroinflammation. Alternatively, neuroinflammation and chronic stress (19) can increase tryptophan 2,3-dioxygenase (TDO) expression in the liver and extrahepatic indoleamine 2,3-dioxygenase (IDO) expression. Activation of these enzymes shunts tryptophan through the kynurenine (Kyn) pathway and therefore potentially compromises the synthesis of serotonin (20). In addition, activation of the Kyn pathway can generate neurotoxic Kyn metabolites that accumulate in the brain during both depression and AD (21-25).

Increased IDO expression has been reported in the hippocampus and neocortex of AD patients (26) and is correlated with beta-amyloid (Aβ) plaque load (27), one of the hallmarks of the disease. In addition, AD subjects demonstrate an activation of the Kyn pathway at the periphery, potentially making tryptophan less available for the synthesis of 5-HT (28). Taken together, these findings point to the possible role of tryptophan metabolism in the pathophysiology of AD and AD comorbid depression. Activation of IDO by inflammation can be measured by the ratio of kynurenine to tryptophan (Kyn/Tryptophan). This ratio increases in inflammation, HIV, AD, and cancer (28-30). In the brain, increased Kyn/5-HT ratios have also been used, to measure the relative decrease in the synthesis of 5-HT that is due to the increased metabolism of tryptophan into Kyn (31).

Thus, the present study was carried out to determine whether inflammation-induced activation of the Kyn pathway and its impact on serotonin metabolism accounted for depressive affect, both domain-specific and global cognitive impairment across the AD spectrum, and AD neuropathological features such as cerebrospinal fluid (CSF) biomarkers and brain atrophy. Finally, because Kyn synthesis, Aβ load, and neuroinflammation are interconnected (27-30), we also used mediation and moderated-mediation (32) to see if associations between these factors accounted for depressive affect scores.

## Methods and Materials

### Setting

The present study used Alzheimer’s Disease Neuroimaging Initiative (ADNI) data. ADNI is a multicenter longitudinal study examining clinical, imaging, genetic, and biochemical markers funded by public and private partnerships in part by the National Institute on Aging, pharmaceutical companies, and foundations through the Foundation for the National Institutes of Health.

### Participants

ADNI1 data was obtained from 746 participants including 211 cognitively normal (CN), 359 Mild Cognitive Impairment (MCI) and 176 AD who had metabolite markers of tryptophan metabolism. Data of interest included: 1) demographics; 2) serum, plasma, and CSF biomarkers, including immunologic markers like pro- and anti-inflammatory cytokines, Aβ, and the neurodegeneration marker tau (33); 3) Magnetic Resonance Imaging (MRI) volumetric scans; 4) neuropsychiatric assessments including self-reported affect and activities of daily living; and 5) neuropsychological performance. Participants were clinically diagnosed at every visit based on standardized criteria described in the protocol manual (http://adni.loni.usc.edu/). Participants taking SSRIs, cholinesterase inhibitors, or NMDA antagonists were excluded to prevent confounding effects, as these medication may influence serotonin metabolite values. For this report, Importantly, ADNI1 excluded prospective participants who had GDS scores reflecting major depression (GDS≥6) and a 1-2 year history of major depression.

### Standard protocol approvals, registrations, and patient consents

Written informed consent was obtained from all ADNI participants at their respective sites. Site-specific Institutional Review Boards approved the ADNI protocol.

### Serum, Plasma, and CSF Biomarkers

Kyn, tryptophan, and 5-HT metabolites were assayed in serum using a Biocrates AbsoluteIDQ p180 kit with liquid chromatography/mass spectrometry from the Alzheimer’s Disease Metabolomics Consortium, as described in white papers (http://adni.loni.usc.edu). In a subset of 58 CN, 396 MCI, and 112 AD, inflammatory markers were assayed from plasma sent to Rules-Based Medicine (RBM, Austin, TX, USA) for analysis using a Luminex xMAP multiplex array (Austin, TX, USA). This array examined 49 biomarkers of immunologic activation, as described in **Supplementary Table 1**. CSF Aβ_1-42_, total tau and phosphorylated (P)Tau-181 were analyzed using xMAP Luminex (Innogenetics/Fujirebio AlzBio3 Ghent, Belgium) immunoassay kits. Each analyte has a validation report after independent evaluation by Myriad RBM with a multianalyte panel (Human Discovery MAP version 1.0; Myriad RBM). Quality control information and the detection limits for assays can be found here: http://adni.loni.ucla.edu/wp-ontent/uploads/2010/11/BC Plasma Proteomics Data Primer.pdf For moderation-mediation analyses, Aβ_1-42_ CSF levels were categorized as being Aβ negative (Aβ-, >192 pg/mL) or positive (Aβ+, ≤192 pg/mL) to reflect clinically meaningful amyloid load for AD (34).

### Apolipoprotein E (APOE) Haplotype

The ADNI Biomarker Core at the University of Pennsylvania conducted APOE s4 haplotyping. We characterized participants as having zero vs. one or two APOE4 alleles.

### MRI Acquisition and Pre-Processing

T1-weighted MR volumetry scans [1.25×1.25×1.25mm] were acquired from 1.5T units within 10-14 days of the screening visit, following a back-to-back 3D magnetization prepared rapid gradient echo (MP-RAGE) scanning protocol described elsewhere (35). Images were pre-processed using FreeSurfer 4.3 (36). As described previously (37), this software corrects for motion, deskulls, bias corrects, segments, and parcellates gray and white matter into labeled areas. Mean gray matter volume was derived from subcortical and archiocortical regions of interest (ROIs), chosen a priori for their relevance to major depression and associations with kynurenine metabolites (38-40). ROIs included: hippocampus, amygdala, caudate, and putamen. Several frontal and cingulate regions were also chosen based on prior studies (41), including Superior Frontal Gyrus, Caudal Anterior Cingulate Gyrus, Medial Orbital Frontal Gyrus, Lateral Orbital Frontal Gyrus, Caudal Middle Frontal Gyrus, and Rostral Middle Frontal Gyrus. In these neocortical areas, we examined cortical thickness (CT) instead of volume. CT is typically a more sensitive index of gray matter pathology in participants with AD risk (42) or who have AD (43).

### Neuropsychological Assessments

All subjects underwent clinical and neuropsychological assessment at the time of scan acquisition. Global tests included Clinical Dementia Rating sum of boxes (CDR-sob), Mini-Mental State Examination (MMSE), AD Assessment Schedule - Cognition 11 (ADAS-Cog). Memory assessments included the Rey Auditory Verbal Learning Test (RAVLT) and a composite memory factor (44). A composite executive function score (45) was also used.

### Neuropsychiatric Assessment

The primary outcome measure of depressive symptoms was the Geriatric Depression Scale, or GDS (46). One sub-score of the GDS asked whether or not a participant felt like they have more memory problems than others. This sub-score was not included in the GDS total due to the high frequency of memory complaints in ADNI. The 12-item Neuropsychiatric Inventory Questionnaire (NPI-Q) and a sub-score examining Apathy/Anxiety were assessed (47). The Functional Assessment Questionnaire (FAQ) measured the ability to carry out 10 activities of daily living according to dependence on a caregiver (48). Higher scores indicated more severe neuropsychiatric symptoms or greater functional impairment.

## Statistical Analysis

All analyses were conducted using SPSS 23 (IBM Corp., Armonk, NY). ANOVA and follow-up LSD tests examined differences in metabolite, cognitive, and other outcomes among CN, MCI, and AD subjects (see **Table 1**). Logistic regression analyses were performed to see if Kyn/5-HT or Kyn/Tryptophan predicted cognitive status (i.e. MCI or AD diagnosis). Subsequently, linear mixed effects models tested the main effects of a Kyn/5-HT or Kyn/Tryptophan ratio on outcomes of interest. Covariates included age at baseline and sex, as well as education for cognitive and affective measures. For subcortical brain volumes, total intracranial volume was also used as a covariate to correct for whole brain size. Other outcomes included: peripheral inflammatory markers (see **Supplementary Table 1**); neuropsychiatric stability; neuropsychological performance; CSF AD biomarkers including Aβ_1-42_, total tau and PTau-181; and subcortical and cortical ROIs.

**Table 1.** Demographics and Sample Characteristics

|  | CN | MCI | AD |
| --- | --- | --- | --- |
| Age | 75.1 ± 5.77 | 74.7 ± 7.40 | 74.80 ± 8.08 |
| Gender (% Male) | 51.7 | 64.6 | 58 |
| Education | 15.7 ± 2.78 | 15.6 ± 3.03 | 15.09 ± 3.20 |
| BMI | 26.89 ± 3.84 | 26.01 ± 3.84 | 25.54 ± 3.84 |
| APOE4 (% carriers) | 8.6<sup>a</sup> | 53.3<sup>b</sup> | 67.9<sup>c</sup> |
| Tryptophan | 71.6 ± 13.4<sup>a</sup> | 71.8 ± 16.00<sup>b</sup> | 72.55 ± 14.66<sup>b</sup> |
| Kyn | 3.33 ± 0.94<sup>a</sup> | 3.15 ± 1.01<sup>a</sup> | 3.31 ± 1.11<sup>a</sup> |
| 5-HT | 0.675 ± 0.37<sup>a</sup> | 0.535 ± 0.340<sup>b</sup> | 0.445 ± 0.407<sup>b</sup> |
| Kyn/5-HT | 9.19 ± 12.60<sup>a</sup> | 13.1 ± 18.8<sup>b</sup> | 23.4 ± 29.4<sup>c</sup> |
| Kyn/Tryptophan | 0.048 ± 0.015<sup>a</sup> | 0.045 ± 0.016<sup>a</sup> | 0.047 ± 0.019<sup>a</sup> |
| CDR-sob | 0.026 ± 0.11<sup>a</sup> | 1.60 ± 0.888<sup>b</sup> | 4.32 ± 1.56<sup>c</sup> |
| MMSE | 28.9 ± 1.15<sup>a</sup> | 27.0 ± 1.78<sup>b</sup> | 23.6 ± 1.91<sup>c</sup> |
| ADAS-cog11 | 6.25 ± 2.79<sup>a</sup> | 11.5 ± 4.42<sup>b</sup> | 18.3 ± 6.42<sup>c</sup> |
| ADNI Memory Factor | 0.87 ± 0.46<sup>a</sup> | -0.086 ± 0.586<sup>b</sup> | -0.814 ± 0.547<sup>c</sup> |
| ADNI EF Factor | 0.71 ± 0.58<sup>a</sup> | -0.043 ± 0.783<sup>b</sup> | -0.934 ± 0.816<sup>c</sup> |
| GDS Total | 0.86 ± 1.23<sup>a</sup> | 1.58 ± 1.37<sup>b</sup> | 1.72 ± 1.36<sup>b</sup> |
| FAQ Total | 0.05 ± 0.22<sup>a</sup> | 3.82 ± 4.46<sup>b</sup> | 12.53 ± 6.71<sup>c</sup> |
| NPI-Q Total | 0.28 ± 0.72<sup>a</sup> | 1.85 ± 2.67<sup>b</sup> | 3.39 ± 3.31<sup>c</sup> |
| Aβ<sub>1-42</sub> (pg/mL) | 251 ± 21.1<sup>a</sup> | 168 ± 55.6<sup>b</sup> | 144 ± 40.8<sup>c</sup> |
| Total Tau (pg/mL) | 63.6 ± 21.8<sup>a</sup> | 97.4 ± 58.1<sup>b</sup> | 119 ± 56.8<sup>c</sup> |
| P-Tau (pg/mL) | 21.1 ± 8.43<sup>a</sup> | 35.5 ± 18.8<sup>b</sup> | 41.8 ± 19.7<sup>c</sup> |

Numbers represent frequency or unadjusted mean ± SD. Superscript letters indicate if a given value for a clinical group is significantly different from other clinical groups. For example, CDR-sob differs between each clinical group, while 5-HT only differs between the CN versus MCI and AD groups. Aβ = beta-amyloid; AD = Alzheimer's disease; ADAS-Cog = Alzheimer's Disease Assessment Scale-cognition 11; APOE4 = apolipoprotein E4; CDR-sob=Clinical Dementia Rating – sum of boxes; CN = Cognitively normal; EF = executive function; GDS = Geriatric Depression Scale; FAQ=Functional Assessment Questionnaire; Kyn = Kynurenine; MCI = Mild Cognitive Impairment; MMSE = Mini-Mental State Exam; NPI-Q=Neuropsychiatric Inventory Questionnaire.

To correct for type 1 error, omnibus MANCOVA testing was used for a given family of outcome variables (e.g., neuropsychological tests). If the omnibus was significant, all follow-up tests were judged at p<.05 because the family-wise error rate stays below Alpha of .05 (50). When the omnibus was non-significant, a stricter Holm-Bonferroni correction (49) was used. This closed test procedure maintains a family-wise Alpha = 0.05 by requiring unadjusted P values of 0.05 divided by x, x being the number of null hypotheses tested. For four cognitive tests, for example, P values of .0125, .025, .0375, and .050 would be successively needed when testing outcomes in the closed set.

Finally, it was of interest to conduct mediation and moderated mediation analyses using the PROCESS macro (32). The objective was to test if immunologic or AD biomarkers accounted for significant associations between Kyn/5-HT and GDS scores. Kyn/Tryptophan was not considered because it was not related to GDS scores. First, to constrain type 1 error, all immunologic mediators were entered and then backwards selection used to retain biomarkers at p<.05. In turn, Kyn/5-HT was regressed onto each selected inflammatory marker, yielding a beta coefficient (e.g., path A). The inflammatory marker was separately regressed onto depression scores (i.e., GDS), yielding a second beta coefficient (e.g., path B). The indirect effect was estimated as the product between the two beta coefficients. The direct effect (path C) was estimated by regressing Kyn/5-HT against GDS scores. The size of the mediation effect was estimated using the variance percentage attributed to the complete model explained by the mediator (51). For moderated mediation, we exclusively focused on Aβ because protein oligomers can influence neuroinflammation and tryptophan metabolism (13, 27-30). Aβ status (Aβ- vs. Aβ+) was tested as a moderator of path B and path C. Covariates in the models included age and gender.

## Results

### Data Summary

Clinical, demographic, and other data and differences among CN, MCI, or AD participants are presented in **Table 1**. As expected in this ADNI sub-sample, there were step-wise declines in global cognition, memory, executive function, amyloid and tau markers, and being APOE4 positive. While ADNI1 did not recruit participants with GDS scores in the major depression range, MCI and AD nonetheless had mild depressive symptoms and more neuropsychiatric disturbances vs. CN.

For CSF metabolites of interest, tryptophan [F=4.29, p=.014] and 5-HT [F=9.27, p<.001] levels were higher in CN participants than MCI or AD. There was also a marked dose-response difference in Kyn/5-HT between CN, MCI, and AD [F=10.93, p<.001]. No differences were noted for Kyn or the Kyn/Tryptophan ratio, by contrast. For peripheral immune biomarkers, **Supplementary Table 1** shows the mean, standard deviation, unit of measurement, and percentage of missingness due to values being below the detection threshold. Many of these variables were log transformed to achieve normality for use in parametric tests.

### Risk of Cognitive Impairment

A higher Kyn/Tryptophan ratio was not associated with baseline clinical diagnosis. A higher Kyn/5-HT ratio, however, was significantly associated with being diagnosed as MCI or AD versus CN [F=26.0, P<0.001], but not MCI conversion to AD. Logistic regression models indicated that per point increase in the Kyn/5-HT ratio, risk doubled for having MCI or AD [OR=1.953, p<0.001, Wald=10.18].

### Peripheral Immunologic Biomarkers

Linear mixed effect models tested whether Kyn/5-HT or Kyn/Tryptophan ratios were related to pro- and anti-inflammatory immune markers available in an ADNI multiplex serum panel (see **Supplemental Table 1**). While analyses were done for all markers, caution is warranted for interpreting markers with 25% or greater missingness. Such markers were not included in mediation or moderated mediation analyses with GDS.

For the Kyn/Tryptophan ratio, a significant multivariate omnibus [F=10.3, P< 0.001] allowed for follow-up linear mixed model tests at a family-wise error rate of p<.05 (50). As noted in **Supplementary Table 2**, a higher Kyn/Tryptophan ratio was related to higher levels of most peripheral immune biomarkers.

Representative associations are depicted for IL-1ra [F=21.8, p<.001] (**Figure 1A**), IL-12p40 [F=57.1, p<.001] (**Figure 1B**), and IL-18 [F=72.9, p<.001] (**Figure 1C**). Likewise, an omnibus for Kyn/5-HT [F= 2.09, p< 0.001] and linear mixed models showed that a higher Kyn/5-HT ratio was related to higher levels of many multiplex biomarkers (**Supplementary Table 3**), though fewer in number than for the Kyn/Tryptophan ratio. Representative associations are shown for IL-1ra [F=12.9, (β=.428±.121, P<0.001] (**Figure 1D**), IL-12p40 [F=9.31, β=.0007±.0002, p<0.001] (**Figure 1E**), and IL-18 [F=9.99, β=.718±.241, P<0.001] (**Figure 1F**).

**Figure 1.**
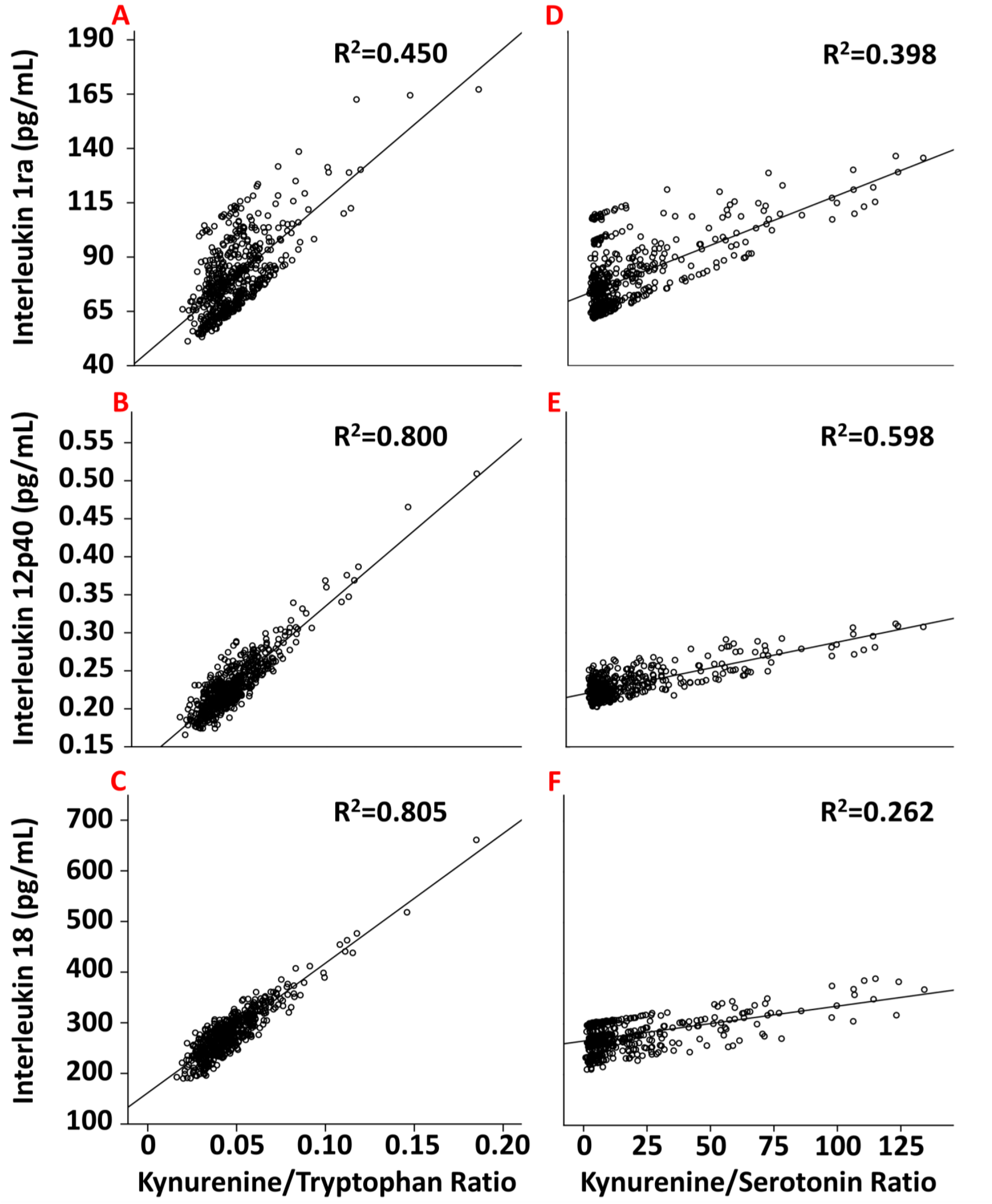
The relationship between Kyn/Tryptophan (A-C, left column) or Kyn/5-HT (D-F, right column) ratios with peripheral immune markers, including IL-1ra; B) IL-12p40; and C) IL-18. R^2^ is the proportion of variance explained by a given ratio.

### Neuropsychiatric Assessments

We next examined Kyn ratios with affect and quality of life outcomes. For Kyn/Tryptophan, the multivariate omnibus was marginally significant [F=2.34, p=.054]. After Holm-Bonferroni correction, higher Kyn/Tryptophan levels were only associated with worse FAQ total scores [F=8.31, β=40.1±13.9, p=.004], indicating less quality of life due to disability. The beta coefficient here is large because the analyte ratio is in the decimal range.

For Kyn/5-HT, the multivariate omnibus was significant [F=3.06, p=.016]. Relationships are noted in **Figure 2** for: **A)** GDS Total; **B)** NPI-Q Total; **C)** FAQ Total; and **D)** the NPI-Q Anxiety sub-score. Subjects with higher Kyn/5-HT ratios had worse scores for the GDS [F=6.28, β=.006±.002, p=012], NPI-Q [F=15.5, β=.016±.004, p<.001], FAQ total [F=42.10, β=.062±.011, p<.001], and NPI-Q anxiety sub-score [F=16.60, β=.002±.001, p<.001].

**Figure 2.**
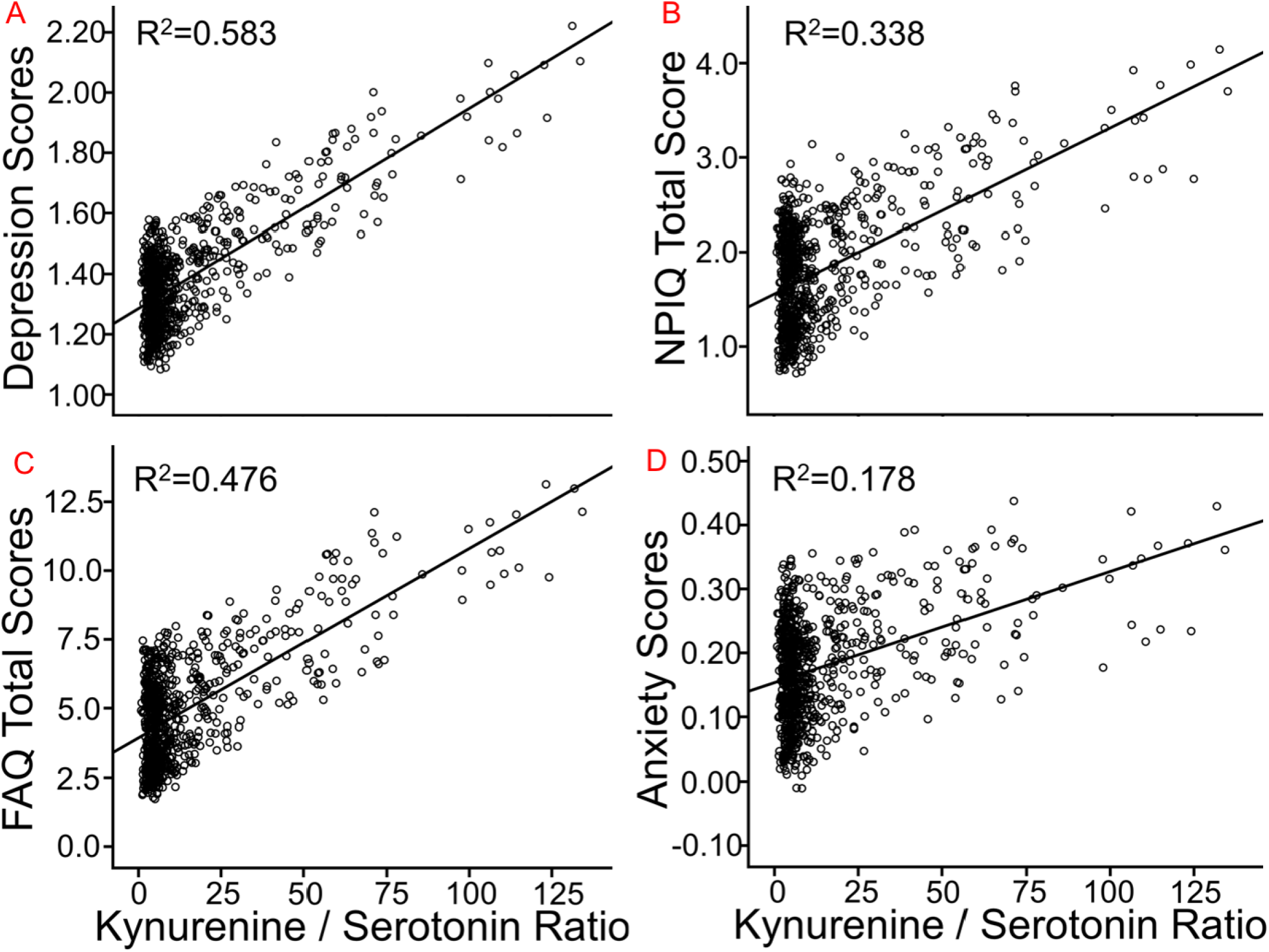
The relationship between Kyn/5-HT ratios and neuropsychiatric assessments, such as: A) GDS total score; B) NPI-Q total score; C) FAQ total score; and D) the anxiety NPI-Q sub-score. R^2^ is the proportion of variance explained by the Kyn/5-HT ratio. FAQ=Functional Assessment Questionnaire; GDS=Geriatric Depression Scale; NPI-Q=Neuropsychiatric Inventory Questionnaire. Note that the depression and anxiety scores respectively represent the GDS and a sub-scale of the NPI-Q. Note also that total GDS score is lower than usual because of removing a question regarding memory concerns, and that several subjects were excluded who took anti-depressive medication.

### Neuropsychological Assessments: Memory and Executive Function

For the Kyn/Tryptophan ratio, the multivariate omnibus was marginally significant. Note again that large beta values are due to the decimal range of the Kyn/Tryptophan ratio. After Holm-Bonferroni correction, a higher ratio corresponded to worse immediate memory on RAVLT Trials 1-5 [F=6.16, β=-60.2±24.3, p=.013] and short delay memory [F=8.26, β=-16.00±5.57, p=.004], as well as a higher percentage of items forgotten during long delay [F=4.32, β=152±73, p=.038]. A higher ratio was also linked to lower Z-scores for the memory factor [F=5.09, β=-4.00±1.77, p=.024].

For Kyn/5-HT, among neuropsychological indices, the multivariate omnibus [F= 5.25, β= 0.002], followed by linear mixed models, indicated that higher Kyn/5-HT ratios were related to worse performance on RAVLT Trials 1-5 (F=14.9, β=-.075±.019, p<0.001) and RAVLT short delay (F=11.5, β=-.016±.004, p<0.001), the memory factor (F=20.4, β=-.006±.001, p<0.001) (**Figure 3A**), and the executive function factor (P<0.001, β=- .006±.002, F=13.9) (**Figure 3B**).

**Figure 3.**
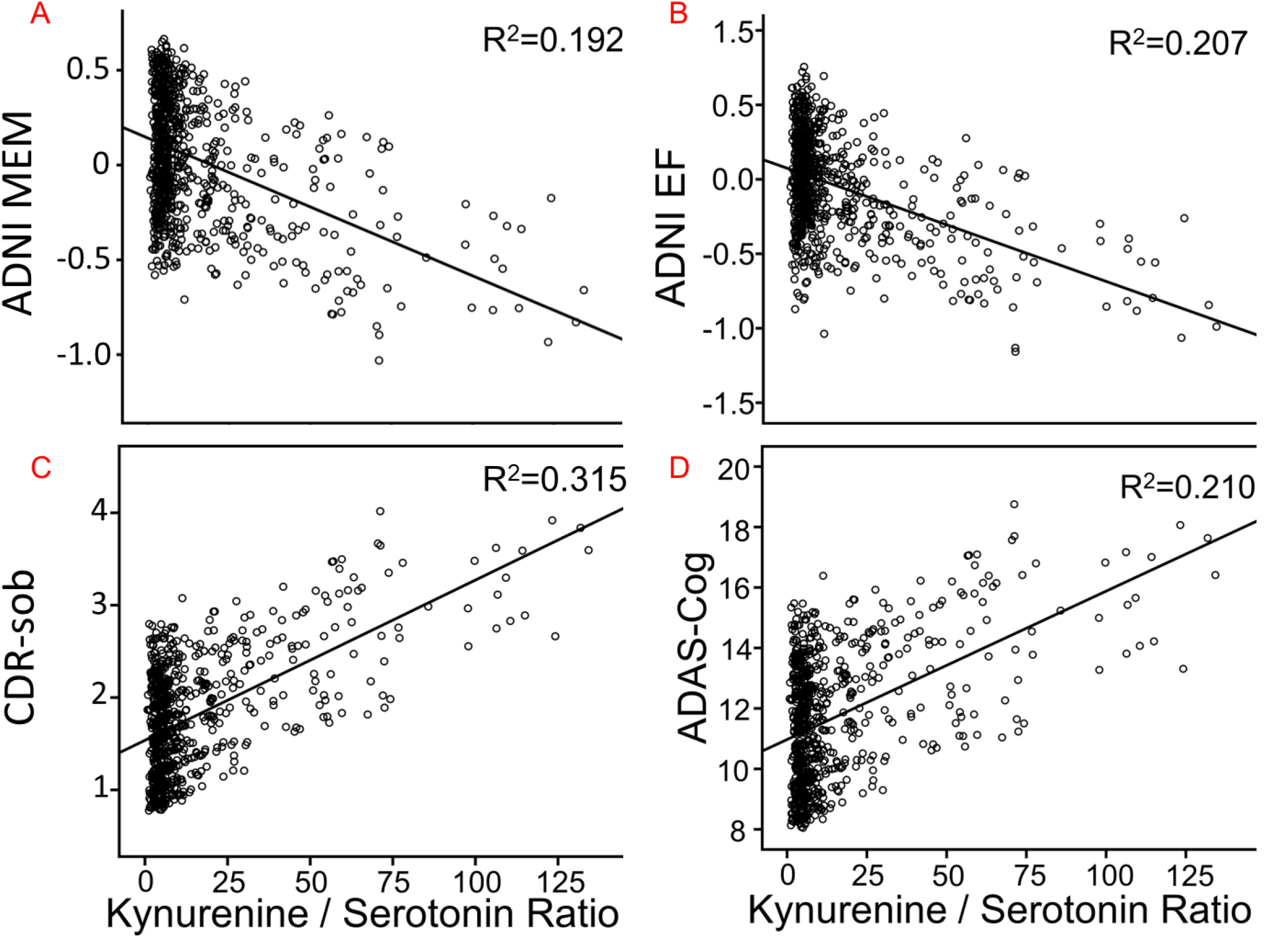
The relationship between Kyn/5-HT ratios and neuropsychological assessments, including: A) ADNI-MEM; B) ADNI-EF; C) CDR-sob; D) ADAS-Cog. R^2^ is the proportion of variance explained by the Kyn/5-HT ratio. ADAS-cog=Alzheimer’s Disease Assessment Scale-cognition 11; CDR-sob=Clinical Dementia Rating - sum of boxes; EF=Executive function factor; MEM=Memory factor.

### Neuropsychological Assessments: Global Domains

For Kyn/Tryptophan, the multivariate omnibus was non-significant and a result with CDR-sob [F=4.391, p=.036] did not survive Holm-Bonferroni correction.

For Kyn/5-HT, a significant omnibus [F=8.70, P=0.001] and follow-up linear mixed models showed that higher Kyn/5-HT ratios were related to worse cognition on CDR-sob (F= 25.0, β=.015±.003, p<0.001) (**Figure 3C**), MMSE (F=14.0, β=-.016±.004, p<0.001), and ADAS-cog (F=14.7, β=.041±.011, p<0.001) (**Figure 3D**).

### CSF Amyloid and Tau

The AD biomarkers of T-Tau, PTau-181 and Aβ_1-42_ were next evaluated. Higher Kyn/Tryptophan ratios were not associated with these indices. By contrast, a multivariate omnibus [F=4.77, p=.003] for Kyn/5-HT and follow-up tests indicated that a higher ratio was associated with lower CSF Aβ_1-42_ (F=11.74, β=-.353±.103, P=0.001) and higher CSF T-Tau (F=3.87, β=.237±.120, p=.05) but not PTau-181, corresponding to increased amyloid and total tau deposition in brain parenchyma.

### Regional Grey Matter Volume

Next, subcortical, hippocampus, and neocortical ROIs implicated in past studies of depression and kynurenine metabolites were investigated. As indicated in **Table 2**, higher Kyn/Tryptophan ratios were not significantly related to gray matter atrophy in any region, though there was a marginal negative association with hippocampus. By contrast, higher Kyn/5-HT corresponded to less hippocampal volume and thinner cingulate and frontal areas chosen a priori.

**Table 2.** MRI Analyses

| Ratio | Gray Matter Index | Brain Region | F Value | $\beta$ Estimate | Standard Error | p value |
| --- | --- | --- | --- | --- | --- | --- |
| Kyn/5-HT | Sub-Cortex and Archiocortex Volumes | **Hippocampus** | **8.70** | **-5.30** | **1.80** | **.003** |
|  |  | Putamen | 0.43 | -1.51 | 2.30 | .511 |
|  |  | Caudate | 0.09 | -0.54 | 1.84 | .769 |
|  |  | Amygdala | 0.41 | -0.46 | 0.71 | .524 |
|  | Neocortex Cortical Thickness | **Superior Frontal Gyrus** | **12.1** | **-1.4E-3** | **4.0E-4** | **.001** |
|  |  | **Caudal Anterior Cingulate Gyrus** | **6.34** | **-1.3E-3** | **5.2E-4** | **.012** |
|  |  | **Medial Orbital Frontal Gyrus** | **12.1** | **-1.3E-3** | **3.7E-4** | **.001** |
|  |  | **Lateral Orbital Frontal Gyrus** | **10.1** | **-1.2E-3** | **3.7E-4** | **.002** |
|  |  | **Caudal Middle Frontal Gyrus** | **8.04** | **-1.1E-3** | **4.0E-4** | **.005** |
|  |  | **Rostral Middle Frontal Gyrus** | **9.16** | **-1.0E-3** | **3.3E-4** | **.003** |
| Kyn/TRP | Sub-Cortex and Archiocortex Volumes | Hippocampus | 3.40 | -4383 | 2379 | .066 |
|  |  | Putamen | 0.18 | -1279 | 3033 | .673 |
|  |  | Caudate | 0.05 | -545 | 2429 | .823 |
|  |  | Amygdala | 0.60 | -730 | 942 | .439 |
|  | Neocortex Cortical Thickness | Superior Frontal Gyrus | 0.95 | -0.52 | 0.53 | .330 |
|  |  | Caudal Anterior Cingulate Gyrus | 1.00 | 0.69 | 0.69 | .318 |
|  |  | Medial Orbital Frontal Gyrus | 1.10 | -0.52 | 0.50 | .295 |
|  |  | Lateral Orbital Frontal Gyrus | 0.11 | -0.16 | 0.49 | .738 |
|  |  | Caudal Middle Frontal Gyrus | 0.01 | 0.48 | 0.54 | .929 |
|  |  | Rostral Middle Frontal Gyrus | 0.52 | -0.31 | 0.44 | .472 |

Brain regions implicated in Region of Interest analyses examining the relationship between either Kyn/5-HT or Kyn/TRP ratios and less gray matter in sub-cortical volumes or thickness of the neocortex. Kyn = kynurenine. TRP = tryptophan. Please note that Beta values are large for Kyn/TRP because values are in the decimal range.

### Moderation-Mediation Analyses of Negative Affect

Given that Kyn/5-HT but not Kyn/Tryptophan was correlated with negative affect, the following analyses were restricted to Kyn/5-HT. Mediation and moderated-mediation models determined: 1) which peripheral immunologic markers significantly mediated the Kyn/5-HT relationship with depressive affect scores (i.e., GDS); and 2) if clinically significant Aβ load (i.e., Aβ+ vs. Aβ-) specific to AD (34) acted as a moderator, modifying how immunologic markers were related to GDS. Among all participants, the following markers significantly predicted GDS total scores after error correction: Complement 3 (C3), eotaxin 1 (EO-1), TNF-related apoptosis-inducing ligand receptor (TRAIL), and vascular endothelial growth factor (VEGF). These results were superseded by significant moderation through Aβ load (all p’s<.01 to .05). Specifically, as shown in **Figure 4**, Aβ- participants showed higher GDS scores when VEGF levels were higher and C3 levels were lower. For Aβ+ participants through full mediation, by contrast, higher C3 drove the association between higher Kyn/5-HT and worse GDS scores.

**Figure 4.**
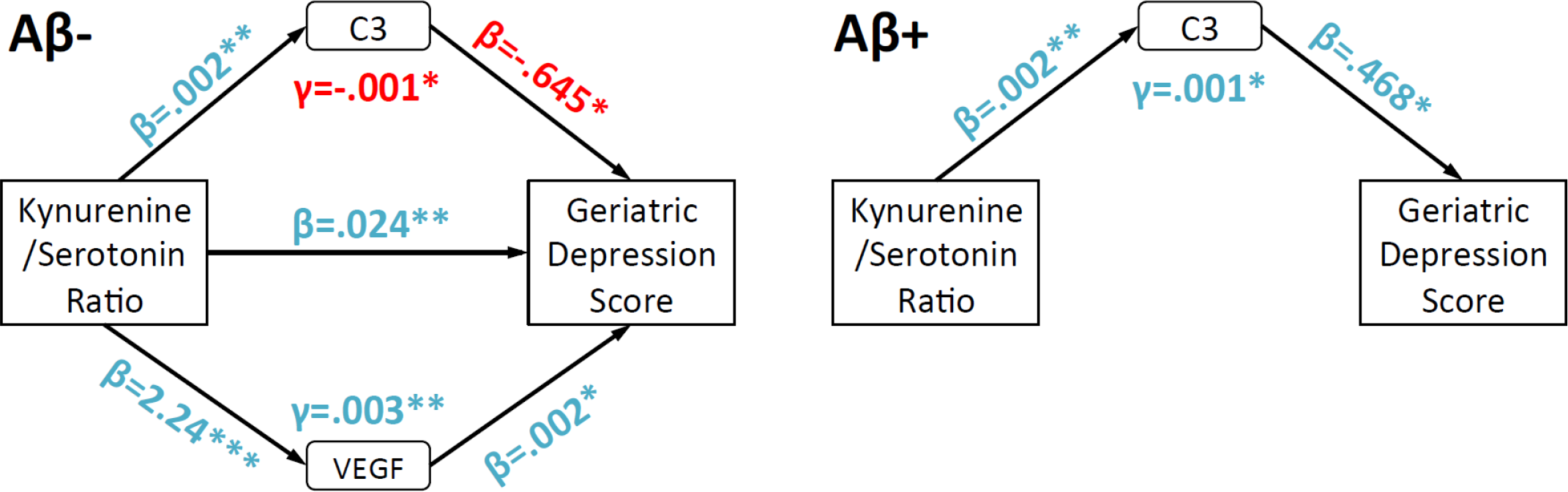
Path diagrams highlighting how immunologic factors and Aβ status modify Kyn/5-HT and Geriatric Depression Scale (GDS) total score associations. Separate path diagrams for Aβ- and Aβ+ are used to illustrate how Aβ status led to different patterns of mediation by immunologic factors. “Blue” and “red” colors respectively highlight positive or negative associations between variables. For clarity, only C3 and VEGF are displayed because they were significant mediators for Aβ- and/or Aβ+. Covariates included age and gender. *,**=p<.05, .01.

## Discussion

Overall, the results suggest that shifted tryptophan metabolism toward the kynurenine pathway is related to not only more self-reported negative affect and behavioral disturbances, but also clinical risk and cognitive impairment, more amyloid and tau deposition, and gray matter atrophy in AD- and depression-sensitive regions. Curiously, for the classic Kyn/Tryptophan ratio, higher values did not correspond to negative affect, but as predicted were strongly associated with peripheral immunologic markers. Higher Kyn/Tryptophan was also strongly related to impaired memory. By contrast, for the first time in a large human sample, we found that a Kyn/5-HT ratio showed immunologic ties similar to Kyn/Tryptophan, but also correlated with negative affect, global cognition and both memory and executive function, AD biomarkers, and regional gray matter in key areas related to emotion regulation and memory. Furthermore, Aβ status modified how Kyn/5-HT was linked to depressive affect scores, with results suggesting that the complement system fully accounted for this linkage in participants who were Aβ+.

Howren et al. found that circulating concentrations of pro-inflammatory cytokines such as IL-1, IL-6, and TNF- α are elevated during depression in humans (52). Similarly, we found elevated IL-1ra among subjects with a higher Kyn/Tryptophan or Kyn/5-HT ratio, which is in agreement with Zunszain et al. (53) who suggested that the production of IL-1 may be impacted by the concentration of Kyn metabolites. This increase in peripheral inflammation may upregulate production of Kyn metabolites and corresponds to our pattern of results.

A primary finding of interest was the strong relationship between a higher Kyn/5-HT ratio with worse global and domain-specific cognition, while Kyn/Tryptophan associations were found exclusively for memory. While progressive memory loss typifies AD, loss of global function and cognition in multiple domains are required for diagnosis. Acute tryptophan deficiency is linked with decreased word recall and impaired memory consolidation (54). The present study shows similar results and supports the theory that less tryptophan is related to worse memory (55), implicating the serotonergic system in these processes. Additionally, only variation in the Kyn/5-HT ratio was strongly related to depression and anxiety measures, which is in agreement with other studies (56). By contrast, Kyn/Tryptophan ratios were not correlated with emotion outcomes. This result was unexpected because Kyn/Tryptophan typically tracks depressive affect in participants with major depression. While a Kyn/5-HT ratio has only been sparingly investigated previously, it was consistently related to pro- and anti-inflammatory cytokines, chemokines, and other immunologic signaling molecules that show increased concentrations among patients with depressive symptoms (52), including IL-6 and IL-1.

It is also well known that endotoxin-induced sickness behavior consists of energy-conserving, withdrawal- oriented behaviors driven by neuroinflammation (57). Indeed, inflammatory and neuroendocrine markers can mediate affective symptoms in rodents (58), rhesus macaques (59), and humans (24). We found that higher Kyn/5-HT was related to higher levels of peripheral C3, EO-1, TRAIL, and VEGF, which in turn were linked with higher GDS scores. Curiously, Aβ status moderated which of these inflammatory factors mediated depressive affect. For Aβ- individuals, VEGF and C3 were related to more and less depressive affect respectively. For Aβ+ adults, strikingly, C3 was instead related to more depressive affect and fully accounted for the Kyn/5-HT and GDS score association. Aβ can inhibit the angiogenic functionality of VEGF and its receptors (60), which may explain why it only arose in Aβ- adults as a relevant partial mediator. Additionally, higher levels of VEGF and Kyn have been observed in individuals with depression and coronary heart disease (61). C3, meanwhile, is a biomarker of early stage activation of the complement system (62), where the CR1 gene underlies C3b receptor synthesis and has been consistently implicated in AD (63). C3 activation optimizes Aβ opsinization and clearance via red blood cells to the liver for degradation, as well as mediates pro-inflammatory responses via the classic and alternate complement systems (64). In our study, for Aβ- participants, more C3 reduced the relationship between higher Kyn/5-HT and GDS scores, which may be due its Aβ clearance properties. For Aβ+, C3 may instead represent chronic neuroinflammation that is correlated with neurodegeneration and cognitive decline in rodent models (65) and humans (66).

Higher Kyn/5-HT, but not Kyn/Tryptophan, was also related to less gray matter in hippocampal, precuneus, and prefrontal cortex volumes or cortical thickness, which complement relationships with affect and cognitive outcomes. It is important to note that Kyn/Tryptophan and regional atrophy have been consistently found, but only in patients with major depression (38-41), which unfortunately were excluded from ADNI1 enrollment. For the first time in aged humans, we also found that Kyn metabolism was correlated with lower Aβ_1-42_ and higher total tau. These data reflect increased amyloid and tau deposition in brain parenchyma and may corroborate how Kyn metabolism impacts amyloid plaque formation (67). It was particularly interesting that clinically significant Aβ (i.e., Aβ+) may influence Kyn metabolism and depressive affect via the complement system. Future work in rodent models should determine if these correlations are meaningful and relevant to common comorbid negative affect in MCI and AD (68, 69).

Several limitations of this study should be highlighted. Foremost, Kyn/Tryptophan was not related to affect or brain volumetry, as is typically shown in this literature. Rather, it was only correlated with immune markers, memory function, and functional independence. One reason may be the ADNI1 sample’s intentionally limited range of GDS scores, the range of which is sub-depressive to mild depression. This contrast may be due to Kyn/Tryptophan representing the degree of neuroinflammation present and how much tryptophan is shunted toward the Kyn pathway, which may not necessarily reflect downstream effects on emotional processes. Another limitation is that many Kyn metabolism end-products, including quinolinic acid or 3-hydroxy-Kyn, have not been measured in ADNI. These metabolites would have provided a more complete picture of the Kyn pathway and which neurotoxic products were associated with neural, cognitive, affective, and AD biomarker outcomes. Several hundred subjects also lacked multiplex data, with many of them being cognitively intact. Thus, caution is warranted for extrapolating the mediation analyses to CN subjects, as most subjects with Kyn metabolites, GDS, and inflammatory marker data were MCI or AD.

Nonetheless, results suggest that Kyn/Tryptophan and especially Kyn/5-HT may be relevant biomarkers of neuroinflammatory pertinent to the pathogenesis of negative affect, cognitive decline, AD biomarkers, and clinical impairment diagosis in AD. In particular, Aβ may be key in changing the complement system from mitigating Kyn related associations with negative affect to exacerbating and fully driving them. Future work will shed light on the role of tryptophan metabolism in functional connectivity among brain networks, as well as further examining associations with peripheral neuroinflammatory markers in relation to affect outcomes.

## Data Availability

All data was downloaded and is accessible to bona fide researchers. Data access instructions are available in the hyperlink we note below.

http://adni.loni.usc.edu/data-samples/access-data/

## Acknowledgements

This study was funded in part by the College of Human Sciences at Iowa State University, a Big Data Brain Initiative grant through the Iowa State University Office of Vice President for Research, NIH grant AG047282, and the Alzheimer’s Association Research Grant to Promote Diversity grant AARGD-17-529552. No funding source had any involvement in the report. Data collection and sharing for this project were funded by the ADNI (National Institutes of Health Grant U01-AG-024904) and Department of Defense ADNI (award number W81XWH-12-2-0012). ADNI is funded by the National Institute on Aging, the National Institute of Biomedical Imaging and Bioengineering, and through generous contributions from the Alzheimer’s Association and the Alzheimer’s Drug Discovery Foundation. The Canadian Institutes of Health Research is providing funds to support ADNI clinical sites in Canada. Private-sector contributions are facilitated by the Foundation for the National Institutes of Health (www.fnih.org). The grantee organization is the Northern California Institute for Research and Education, and the study is coordinated by the Alzheimer’s Disease Cooperative Study at the University of California, San Diego. ADNI data are disseminated by the Laboratory for Neuroimaging at the University of Southern California. The data used in the preparation of this article were obtained from the ADNI database (adni.loni.usc.edu). As such, the investigators within the ADNI contributed to the design and implementation of ADNI and/or provided data but did not participate in analysis or writing of this report. Portions of this report were presented at the Psychoneuroimmunology Research Society in 2017 in Galveston, Texas.

---

**Supplementary Table 1.** Multiplex array of pro- and anti-inflammatory immunologic markers

| Immunologic Marker | Abbreviation | Unit | Mean ± SD | % of Samples Below LDD |
| --- | --- | --- | --- | --- |
| C-Reactive Protein | CRP | ug/mL | 3.23 ± 6.63 | 0% |
| CD40 Antigen | CD40 | ng/mL | 0.790 ± 0.240 | 0% |
| CD40 Ligand | CD40-L | ng/mL | 0.460 ± 0.560 | 0.177% |
| Complement C3 | C3 | mg/mL | 1.79 ± 0.340 | 0.00% |
| Eotaxin-1 | Eo-1 | pg/mL | 86.0 ± 48.8 | 0.707% |
| Fibroblast Growth Factor - Basic | FGF-Basic | pg/mL | 149 ± 116 | 35.5% |
| Fibroblast Growth Factor 4 | FGF-4 | pg/mL | 364 ± 620 | 5.83% |
| Granulocyte Colony-Stimulating Factor | G-CSF | pg/mL | 7.01 ± 5.82 | 11.1% |
| Granulocyte-Macrophage Colony-Stimulating Factor | GM-CSF | pg/mL | 16.0 ± 17.6 | 83.2% |
| Growth-Regulated Alpha Protein | GRO-alpha | pg/mL | 612 ± 497 | 0% |
| Hepatocyte Growth Factor | HGF | ng/mL | 4.14 ± 1.72 | 0% |
| Interferon-γ | IFN-γ | pg/mL | 3.25 ± 3.84 | 74.2% |
| IL-1β | IL-1β | pg/mL | 1.12 ± 4.31 | 80.7% |
| Interleukin-1 Receptor Antagonist | IL-1ra | pg/mL | 81.1 ± 58.5 | 12.7% |
| Interleukin-3 | IL-3 | ng/mL | 0.030 ± 0.020 | 9.72% |
| Interleukin-5 | IL-5 | pg/mL | 2.66 ± 2.21 | 22.8% |
| Interleukin-6 | IL-6 | pg/mL | 10.5 ± 123 | 32.2% |
| Interleukin-6 receptor | IL-6r | ng/mL | 30.5 ± 8.77 | 0% |
| Interleukin-7 | IL-7 | pg/mL | 19.5 ± 13.6 | 57.6% |
| Interleukin-8 | IL-8 | pg/mL | 11.5 ± 5.87 | 0.707% |
| Interleukin-10 | IL-10 | pg/mL | 6.91 ± 20.7 | 36.9% |
| Interleukin-12 subunit p40 | IL-12p40 | ng/mL | 0.234 ± 0.110 | 28.4% |
| Interleukin-13 | IL-13 | pg/mL | 42.4 ± 10.9 | 2.30% |
| Interleukin-15 | IL-15 | pg/mL | 0.30 ± 0.17 | 24.0% |
| Interleukin-16 | IL-16 | pg/mL | 376 ± 162 | 0% |
| Interleukin-18 | IL-18 | pg/mL | 279 ± 125 | 0% |
| Interleukin-25 | IL-25 | pg/mL | 8.43 ± 4.91 | 56.0% |
| Macrophage Colony-Stimulating Factor 1 | M-CSF | ng/mL | 0.040 ± 0.010 | 7.24% |
| Macrophage Inflammatory Protein-1α | MIP-1α | pg/mL | 172 ± 49.9 | 0% |
| Macrophage Inflammatory Protein-1β | MIP-1β | pg/mL | 177 ± 302 | 0% |
| Macrophage Inflammatory Protein-3α | MIP-3α | pg/mL | 77.9 ± 122 | 0% |
| Macrophage Migration Inhibitory Factor | MIF | ng/mL | 0.530 ± 0.530 | 0% |
| Matrix Metalloproteinase-1 | MM-1 | ng/mL | 1.69 ± 4.68 | 0.353% |
| Matrix Metalloproteinase-2 | MM-2 | ng/mL | 4157 ± 1272 | 0.180% |
| Matrix Metalloproteinase-3 | MM-3 | ng/mL | 0.150 ± 0.690 | 33.2% |
| Matrix Metalloproteinase-7 | MM-7 | ng/mL | 2.28 ± 7.48 | 0% |
| Matrix Metalloproteinase-9 | MM-9 | ng/mL | 164 ± 104 | 0.353% |
| Matrix Metalloproteinase-9 Total | MM-9T | ng/mL | 251 ± 121 | 0% |
| Matrix Metalloproteinase-10 | MM-10 | ng/mL | 0.060 ± 0.130 | 0% |
| Monocyte Chemotactic Protein 1 | MCP-1 | pg/mL | 180 ± 286 | 0% |
| Monocyte Chemotactic Protein 3 | MCP-3 | pg/mL | 14.5 ± 48.0 | 6.18% |
| Monokine Induced by Gamma Interferon | MIG | pg/mL | 4111 ± 2940 | 0% |
| Platelet-Derived Growth Factor BB | PDGF-BB | pg/mL | 2497 ± 2977 | 1.24% |
| Receptor for Advanced Glycosylation End Products | RAGE | ng/mL | 0.590 ± 0.270 | 0% |
| Stem Cell Factor | SCF | pg/mL | 291 ± 147 | 0% |
| T-Cell-Specific Protein RANTES | RANTES | ng/mL | 1.04 ± 0.40 | 0% |
| Thymus-Expressed Chemokine | TEC | ng/mL | 2.26 ± 0.210 | 0% |
| TNF-Related Apoptosis-Inducing Ligand Receptor | TRAIL | ng/mL | 16.6 ± 7.5 | 0% |
| Tumor Necrosis Factor - $\alpha$ | TNF-α | pg/mL | 8.96 ± 7.89 | 6.89% |
| Vascular Endothelial Growth Factor | VEGF | pg/mL | 659 ± 269 | 0% |

**Supplementary Table 2.**
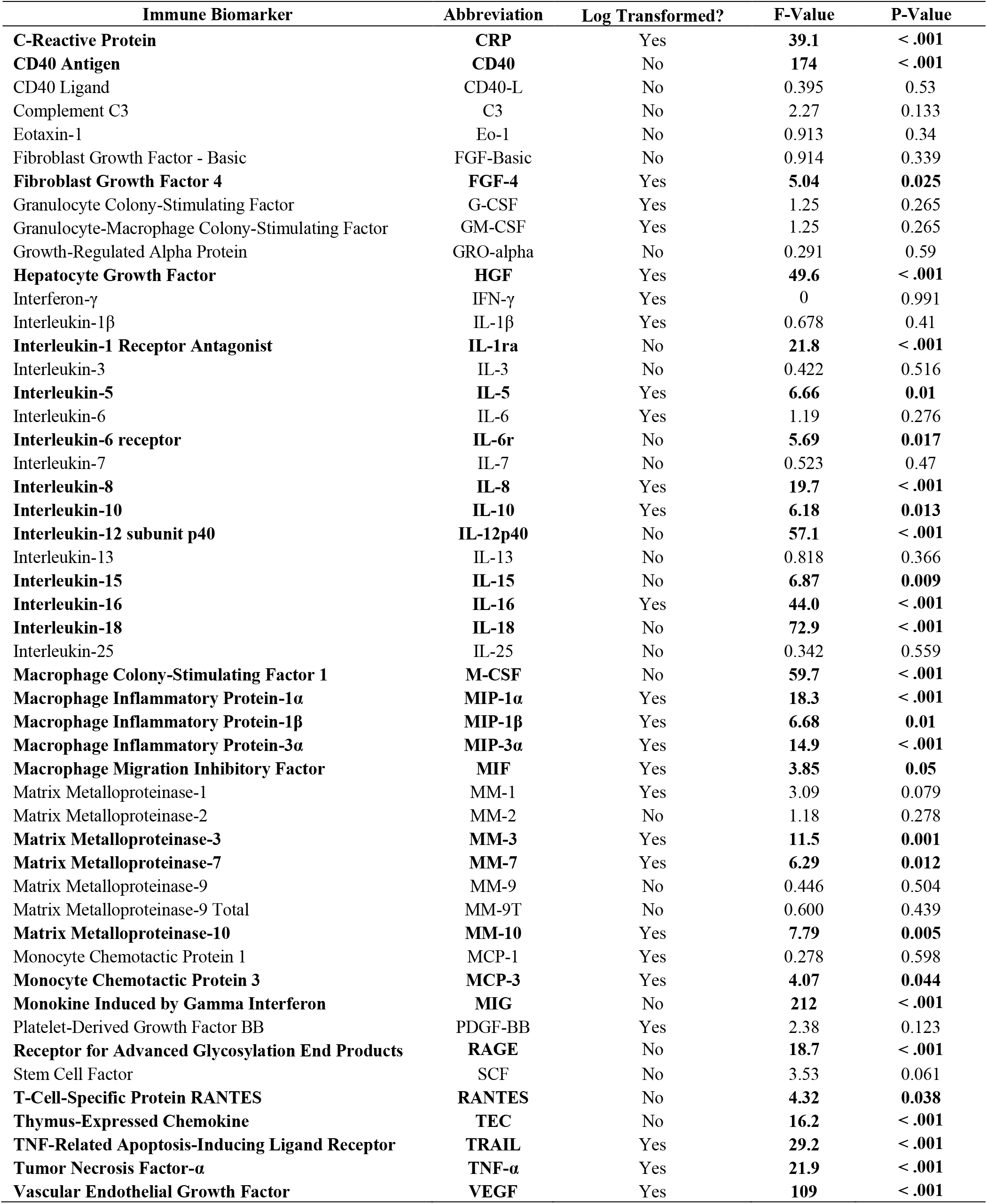
Kyn/Tryptophan and peripheral immunologic biomarker associations

**Supplementary Table 3.**
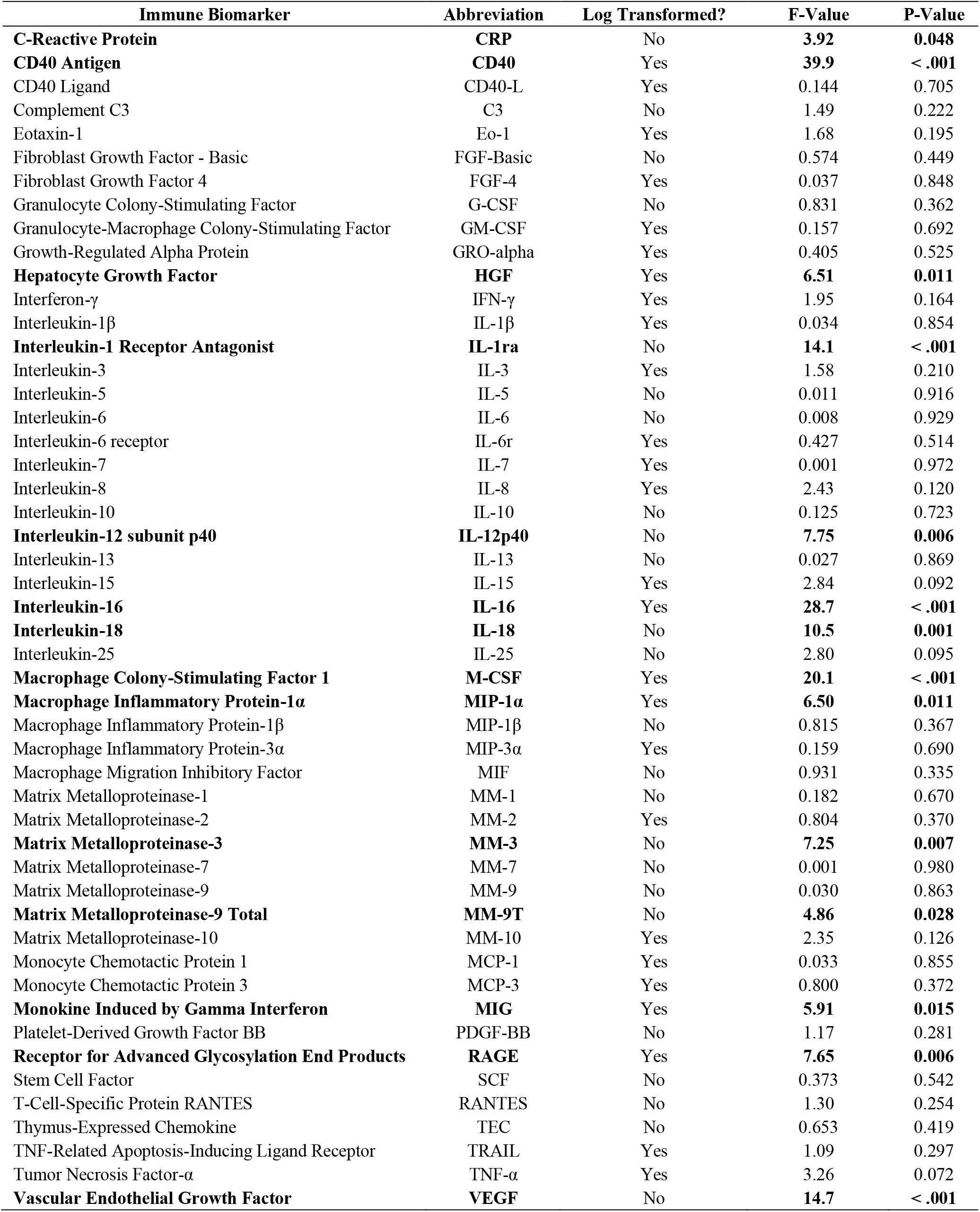
Kyn/5-HT and peripheral immunologic biomarker associations

| Immune Biomarker | Abbreviation | Log Transformed? | F-Value | P-Value |
| --- | --- | --- | --- | --- |
| **C-Reactive Protein** | **CRP** | No | **3.92** | **0.048** |
| **CD40 Antigen** | **CD40** | Yes | **39.9** | **< .001** |
| CD40 Ligand | CD40-L | Yes | 0.144 | 0.705 |
| Complement C3 | C3 | No | 1.49 | 0.222 |
| Eotaxin-1 | Eo-1 | Yes | 1.68 | 0.195 |
| Fibroblast Growth Factor - Basic | FGF-Basic | No | 0.574 | 0.449 |
| Fibroblast Growth Factor 4 | FGF-4 | Yes | 0.037 | 0.848 |
| Granulocyte Colony-Stimulating Factor | G-CSF | No | 0.831 | 0.362 |
| Granulocyte-Macrophage Colony-Stimulating Factor | GM-CSF | Yes | 0.157 | 0.692 |
| Growth-Regulated Alpha Protein | GRO-alpha | Yes | 0.405 | 0.525 |
| **Hepatocyte Growth Factor** | **HGF** | Yes | **6.51** | **0.011** |
| Interferon-γ | IFN-γ | Yes | 1.95 | 0.164 |
| Interleukin-1β | IL-1β | Yes | 0.034 | 0.854 |
| **Interleukin-1 Receptor Antagonist** | **IL-1ra** | No | **14.1** | **< .001** |
| Interleukin-3 | IL-3 | Yes | 1.58 | 0.210 |
| Interleukin-5 | IL-5 | No | 0.011 | 0.916 |
| Interleukin-6 | IL-6 | No | 0.008 | 0.929 |
| Interleukin-6 receptor | IL-6r | Yes | 0.427 | 0.514 |
| Interleukin-7 | IL-7 | Yes | 0.001 | 0.972 |
| Interleukin-8 | IL-8 | Yes | 2.43 | 0.120 |
| Interleukin-10 | IL-10 | No | 0.125 | 0.723 |
| **Interleukin-12 subunit p40** | **IL-12p40** | No | **7.75** | **0.006** |
| Interleukin-13 | IL-13 | No | 0.027 | 0.869 |
| Interleukin-15 | IL-15 | Yes | 2.84 | 0.092 |
| **Interleukin-16** | **IL-16** | Yes | **28.7** | **< .001** |
| **Interleukin-18** | **IL-18** | No | **10.5** | **0.001** |
| Interleukin-25 | IL-25 | No | 2.80 | 0.095 |
| **Macrophage Colony-Stimulating Factor 1** | **M-CSF** | Yes | **20.1** | **< .001** |
| **Macrophage Inflammatory Protein-1α** | **MIP-1α** | Yes | **6.50** | **0.011** |
| Macrophage Inflammatory Protein-1β | MIP-1β | No | 0.815 | 0.367 |
| Macrophage Inflammatory Protein-3α | MIP-3α | Yes | 0.159 | 0.690 |
| Macrophage Migration Inhibitory Factor | MIF | No | 0.931 | 0.335 |
| Matrix Metalloproteinase-1 | MM-1 | No | 0.182 | 0.670 |
| Matrix Metalloproteinase-2 | MM-2 | Yes | 0.804 | 0.370 |
| **Matrix Metalloproteinase-3** | **MM-3** | No | **7.25** | **0.007** |
| Matrix Metalloproteinase-7 | MM-7 | No | 0.001 | 0.980 |
| Matrix Metalloproteinase-9 | MM-9 | No | 0.030 | 0.863 |
| **Matrix Metalloproteinase-9 Total** | **MM-9T** | No | **4.86** | **0.028** |
| Matrix Metalloproteinase-10 | MM-10 | Yes | 2.35 | 0.126 |
| Monocyte Chemotactic Protein 1 | MCP-1 | Yes | 0.033 | 0.855 |
| Monocyte Chemotactic Protein 3 | MCP-3 | Yes | 0.800 | 0.372 |
| **Monokine Induced by Gamma Interferon** | **MIG** | Yes | **5.91** | **0.015** |
| Platelet-Derived Growth Factor BB | PDGF-BB | No | 1.17 | 0.281 |
| **Receptor for Advanced Glycosylation End Products** | **RAGE** | Yes | **7.65** | **0.006** |
| Stem Cell Factor | SCF | No | 0.373 | 0.542 |
| T-Cell-Specific Protein RANTES | RANTES | No | 1.30 | 0.254 |
| Thymus-Expressed Chemokine | TEC | No | 0.653 | 0.419 |
| TNF-Related Apoptosis-Inducing Ligand Receptor | TRAIL | Yes | 1.09 | 0.297 |
| Tumor Necrosis Factor-α | TNF-α | Yes | 3.26 | 0.072 |
| **Vascular Endothelial Growth Factor** | **VEGF** | No | **14.7** | **< .001** |

